# Development of an Interactive Web Dashboard to Facilitate the Reexamination of Pathology Reports for Instances of Underbilling of CPT Codes

**DOI:** 10.1101/2022.11.28.22282835

**Authors:** Jack Greenburg, Yunrui Lu, Shuyang Lu, Uhuru Kamau, Robert Hamilton, Jason Pettus, Sarah Preum, Louis Vaickus, Joshua Levy

## Abstract

Current Procedural Terminology Codes is a numerical coding system used to bill for medical procedures and services and crucially, represents a major reimbursement pathway. Given that Pathology services represent a consequential source of hospital revenue, understanding instances where codes may have been misassigned or underbilled is critical. Several algorithms have been proposed that can identify improperly billed CPT codes in existing datasets of pathology reports. Estimation of the fiscal impacts of these reports requires a coder (i.e., billing staff) to review the original reports and manually code them again. As the re-assignment of codes using machine learning algorithms can be done quickly, the bottleneck in validating these reassignments is in this manual re-coding process, which can prove cumbersome. This work documents the development of a rapidly deployable dashboard for examination of reports that the original coder may have misbilled. Our dashboard features the following main components: 1) a bar plot to show the predicted probabilities for each CPT code, 2) an interpretation plot showing how each word in the report combines to form the overall prediction, 3) a place for the user to input the CPT code they have chosen to assign. This dashboard utilizes the algorithms developed to accurately identify CPT codes to highlight the codes missed by the original coders. In order to demonstrate the function of this web application, we recruited pathologists to utilize it to highlight reports that had codes incorrectly assigned. We expect this application to accelerate the validation of reassigned codes through facilitating rapid review of false positive pathology reports. In the future, we will use this technology to review thousands of past cases in order to estimate the impact of underbilling has on departmental revenue.

## Introduction

The assignment of Current Procedural Terminology (CPT) Codes represents a critical compensatory component of a hospital’s financial system. As the pathology laboratory is responsible for a consequential fraction of services billed, assignment of CPT codes (i.e., coding) via the pathology reporting system is essential for generating hospital revenue. Misbilled codes (i.e., over/underbilling) can cost a hospital significant revenue. The value of identifying misbilled codes has been recognized across many medical subspecialties ^1–7^. Pathology reports contain auditable information describing diagnostic case information and additional background information pertaining to services rendered, including what tests/services had been run for the patient and subjective assignment of case complexity. In order for the hospital to receive compensation for these tests, hospitals employ billing staff (i.e., coders) to read, identify, and assign the CPT codes that dictate what tests/services were performed. Accuracy in this coding process is very important since a failure to report any code represents revenue lost by the hospital. Any system that could serve as a “second check” or “suggestion” would have clear and immediate benefits ^8^.

There are several factors that complicate assignment of CPT codes. Pathology reports do not follow a strict format (i.e., reporting standardized to such a degree that a series of clicks are all that are necessary to both provide a case description and assign codes). In fact, reports may vary drastically from each other (i.e., subjective component) based on the personal style of the signing pathologist, their primary language and where they were trained. Not only does each patient present a unique set of diagnostic/prognostic criteria (which may require ordering many different procedures or stains to hone in on a clear diagnosis), but pathologists may describe similar cases with widely varying lexicon (e.g., certain pathologists may be more verbose than their colleagues or vice versa). An added challenge is that the assignment of primary CPT codes (i.e., CPT 88300-88309), is partially based on the perceived complexity of the case at hand (there do exist guiding principles), meaning that for some codes the coding process relies on a subjective interpretation of report text as it aligns to these criteria.

Difficulties of assigning CPT codes may be ameliorated through the use of natural language processing (NLP), which provides an automated and objective means to parse through reporting text to arrive at a key finding. While many NLP methods require the use of hard-programmed rules (e.g., regular expressions to identify specific words), machine learning methods provide the level of adaptiveness that a complex search task (i.e., identifying the CPT code) requires by learning, so to speak, patterns of text that signify assignment of specific codes ^9^. Many current NLP projects exist to convert free text from pathology reports into structured electronic health record (EHR) information for synoptic case reporting and experiment planning (i.e., a structured search tool for identifying cases to form a study cohort) ^10–15^.

The topic of CPT code assignment remains relatively less explored in comparison. A 2019 paper by JJ Ye began to explore the concept of applying machine learning to assist in coding. Ye concluded that augmenting existing coding pipelines with machine learning technologies may prove beneficial and could potentially help avoid misbilling ^16^.

Our research group is keen to understand the impact of underbilling on lost hospital revenue. We hypothesize that codes predicted by the machine learning model that are higher than the manually assigned codes have a high likelihood of representing erroneous underbilling. However, we are unable to explore this possibility as no publicly available dashboards exist to help facilitate the process of estimating the scope and impact of misbilling. While there do exist dashboards to assist in pathology report analysis, they do not operate within our specific niche ^17^. In this study, we document the development of a web application that assists with interrogating the results from an NLP prediction pipeline and comment on its potential application for identifying misbilling. We imagine this tool would serve as a research resource for others to employ in similar assessments. In a future work, we hope to employ such a dashboard to review thousands of past cases in order to provide information on how much money is lost by the hospital due to mistakes in coding. Further in the future, **the dashboard will be useful tool to easily test the viability of models, accelerating the automation timeline for the assignment of CPT codes (Figure 1)**.

**Figure 1:**
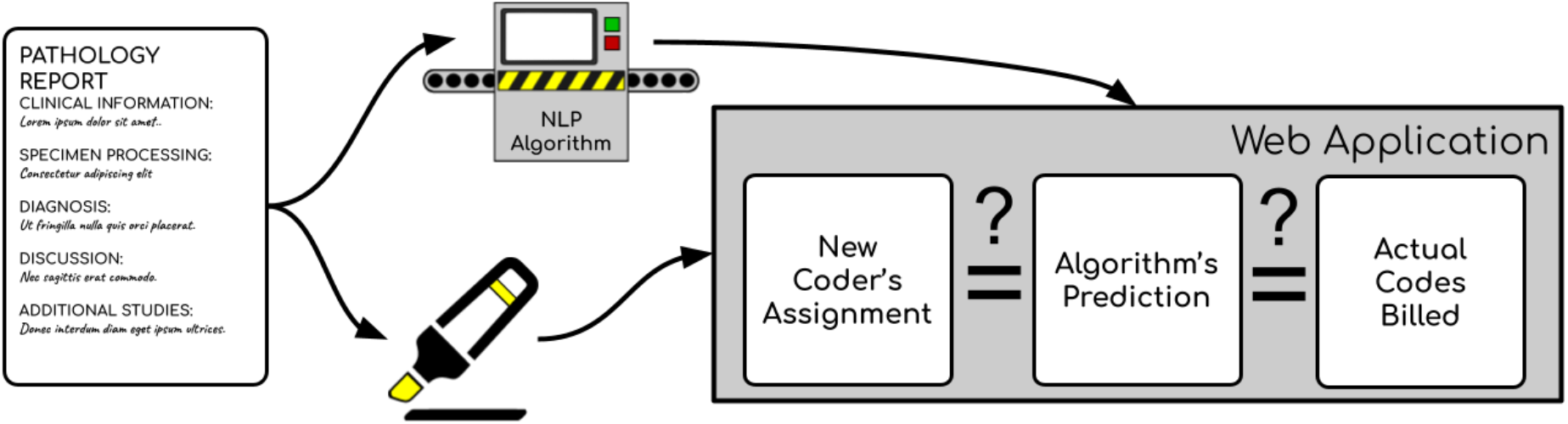
Graphical depiction of key innovations: Pathology report is processed by coder and NLP algorithm; coder reviews candidate underbilled codes to determine if code was actually underbilled

## Methods

### Data Collection

We utilized the data from 93,039 pathology individual pathology reports, as previously documented in a prior study (5 year period) ^18^. Of the data that we had access to, we utilized only the report text and the CPT codes that were identified by the coders. While many CPT codes were assigned to these cases, we trained machine learning models to recognize the 38 most prevalent codes. Most of these are ancillary codes (codes assigned with the click of a button for ordered procedures), while we denoted five such codes as primary codes based on case complexity (CPT 88302, 88304, 88305, 88307, 88309). Data collection and preprocessing follows a prior work. Diagnostic and other reporting subsections were formatted using the spaCy and scikit-learn python packages to generate count matrices (i.e., rows denote reports, columns denote words, elements represent term frequency).

### Model Training and Dataset Partition

We train the models using XGBoost. XGBoost is gradient boosting decision tree library which generates decision trees that are iteratively morphed based on the errors obtained from the previously fit trees ^19^. However, other machine learning models (e.g., BERT or another algorithm) could be substituted with minor respecification of the preprocessing workflow ^20^. We chose XGBoost because it was faster to implement, and was found to be slightly more accurate than BERT in a previous work ^18^. Comparing the accuracy of specific algorithms is outside of the scope of this work as our aim is to document the display outputs that interface with these algorithms.

We trained two separate sets of XGBoost models which can interface with this dashboard. First, we trained a model to differentiate primary CPT codes 88302, 88304, 88305, 88307, and 88309. These codes pertain to varied degrees of complexity for assessment of different conditions. For each set of codes, we trained two versions of each model, one with just the diagnosis section and another with the full report text (i.e., adding other reporting subfields). Next, we trained a second set of models on the 38 most common CPT codes as detailed in our previous work. These models can interrogate CPT codes that correspond to common ancillary tests (e.g., immunostaining), though this was not the primary focus of this work.

### Web Application Development

As potential ambiguity of free text within pathology reports could lead to the erroneous assignment of primary codes on part of the human rater, we would expect these inaccuracies to be well accounted for using our modeling approach. We developed an interactive web application to facilitate the assessment of instances where our model disagreed from the original CPT code assignment. We have included further detail on the design of the dashboard in this section.

#### Web application overview

We designed an interactive display output– designed using the Plotly Dash framework– that interfaces with trained machine learning models and allows domain experts to select specific reports to interrogate (**Figure 2**) ^21^. Coders operate this dashboard by first selecting a code or report of interest through a text box at the bottom of the dashboard which can search for reports with specific code assignments, text present (**Figure 2D**), or whether they were predicted to be false positives (and particularly false positives of higher case complexity) as denoted by the model (**Figure 2C**). The prediction plot on the left indicates the predicted probabilities of specific codes for the report in question (**Figure 2A**). The interface also has a prediction box which allows coders utilizing the dashboard to reassign CPT codes based on presented information (i.e., original code assignment, model prediction, etc.). A key functionality here is the ability to apply filters to limit which reports the dashboard will display to them, for example allowing the user to assess false negatives (**Figure 2C**). In order to facilitate reassessment, this dashboard includes interpretation plots which highlight words associated with select CPT codes to rapidly corroborate findings with established CPT reporting guidelines (linked references to guidelines / code reference). The interpretation plots utilize SHAP (SHapley Additive exPlanations) values to provide additional insight into how the algorithm made its decisions by demonstrating how each specific input word contributes to the overall CPT code prediction ^22^. SHAP values fall under the umbrella of additive feature attribution methods (i.e. methods that assign a numerical importance value to each feature such that all features sum to the prediction value). These values help to unpack the machine learning algorithm black box, and, in our case, they will help to reveal what patterns the algorithms detect in the text.

**Figure 2.**
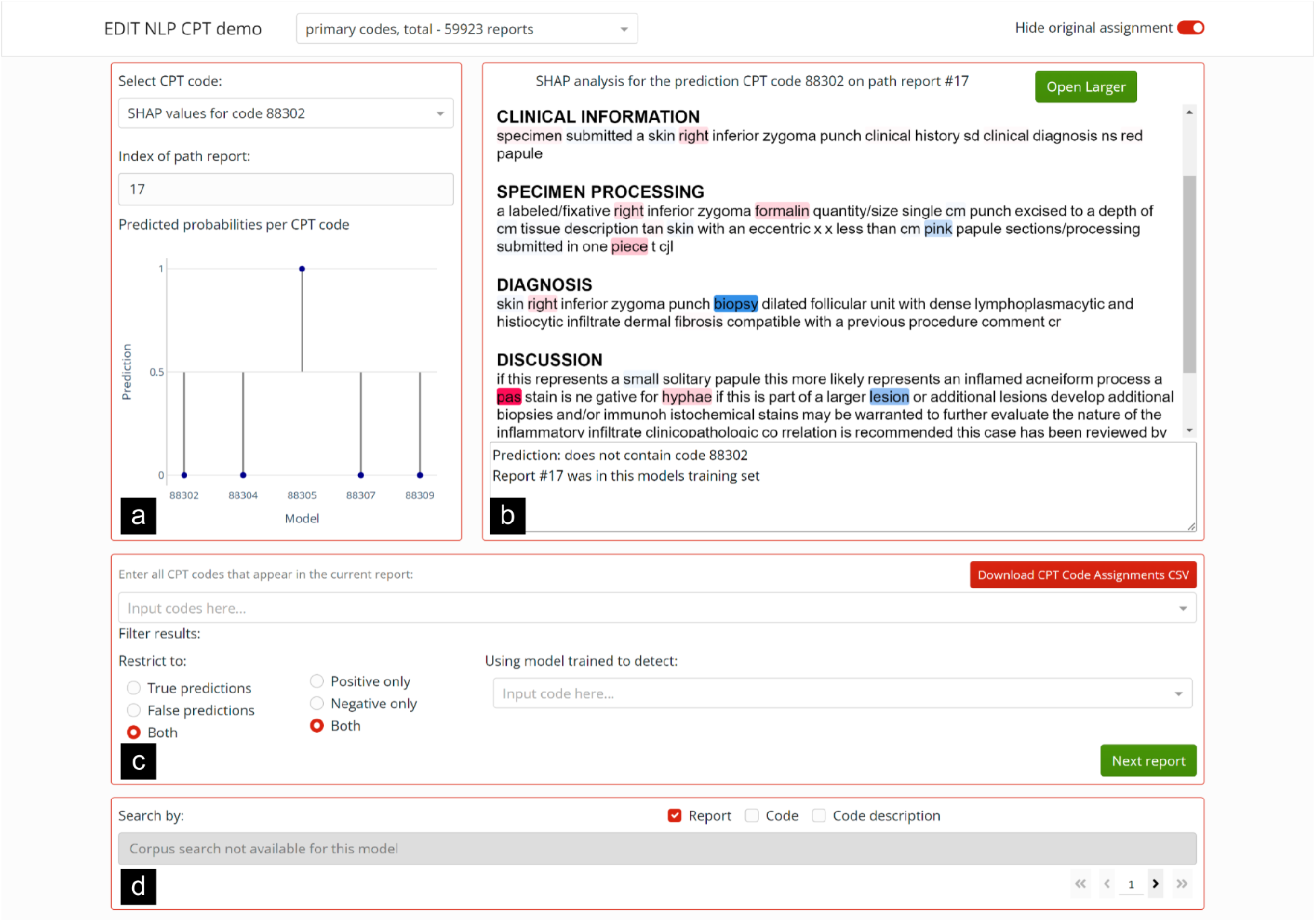
Dashboard overview: a) The bar plot shows the predictions for each CPT code. b) Contains the interpretation plot for a specific report and specific CPT code. c) Where the user selects which reports to display; it is also where the user will input their assignment. d) Search box to search all reports for specific language.

#### Assigning and submitting new CPT codes

The dashboard allows codes to be displayed in the following ways: 1) originally assigned code, 2) reassigned code, and 3) hide code. Display of the original code can be toggled using the *prediction mode* feature that reveals this prediction box and hides the CPT codes previously chosen by the coders, so as not to bias the user inadvertently. Coders can submit new CPT code reassignments, blinded by assignment of the original code but informed by the machine learning derived prediction. The submitted prediction is added to an output file for later analysis (i.e., underbilling). These reassignments can also be used to help train future iterations of these prediction models to reduce further errors/edits.

#### Methods to explore validity of CPT code assignment and underbilling

We have included additional text on how the SHAP interpretation plots can be configured for different CPT codes and how to filter reports to focus on false positives to facilitate exploration of underbilling candidates in the supplementary material (section “Supplementary Methods”).

### Estimating the Potential Impact of Underbilling

Using data tables downloaded from the Medicare Physician Fee Schedule (MPFS) Look-Up Tool (2022), omitting the modifiers “26” and “TC” (incomplete examination and technical component respectively), we recorded the average non-facility prices for CPT codes 88302-88309: 1) 88302– $33.32, 2) 88304– $43.72, 3) 88305– $74.39, 4) 88307– $301.33, and 5) 88309– $457.70 ^23^. We assessed a subset of pathology reports over a five-year span for an estimate of how these primary codes could potentially be reassigned given reexamination. Multiplying the price difference between candidate underbilled reports (e.g., 88307 vs 88305 indicates $301.33-$74.39=$226.94 difference) by the number of reports yields an upper estimate for the potential savings under the assumption that all candidate underbilled reports were truly underbilled. Using our laboratory information system, we pulled and counted all reports over a ten-year span, from January 2011 to December 2021 to extrapolate these findings. This database search identified 749,136 reports containing diagnostic text and including addendums which will be the subject of future inquiry through the developed dashboard.

### Code Availability

The dashboard is freely available and can be adapted to fit a wide variety of models and purposes. We have included the source code and a small set of examples at the following URL:https://github.com/jackgreenburg/cpt_code_app. The software can be installed locally using Python (*pip*) from this GitHub repository. Note that installing web application locally requires installation of Python 3.8 or above. We have also included a small online demo at the following URL:https://edit.cpt.code.demo.levylab.host.dartmouth.edu/ (**user:** edit_user, **password:** qdp_2022).

## Results

### Examples to Illustrate Operation and Usage of the Web App

Someone capable of assigning CPT codes (i.e., a coder) presented with the dashboard will reassign CPT codes via the following process: 1) Select which refined sequence of reports they want to view, 2) View the interpretation plots for each code, 3) Submit the new CPT code assignment. In the following section, we demonstrate several examples of reports analyzed using the web application to provide an idea on the scope of its usage:

#### Example of true positive cases correctly assigned, e.g., CPT 88305

Using the dashboard with the primary code model (i.e., case complexity of examination under a microscope–CPT codes 88302, 88304, 88305, 88307, and 88309) we evaluated whether reports were likely to be improperly billed. In Figure **2**, we demonstrate a simple example of how the SHAP interpretation plot can be employed to assess the validity of the finding. In this example, the primary code model assigned CPT code 88305. The interpretation plot identified words relating to punch biopsies for assignment of the code for a Dermatopathology case. Punch biopsies are commensurate with assignment of code 88305 as compared to more complex excisions in Dermatopathology ^24^. If this specific report was presented to a coder through this dashboard prior to assignment of a CPT code, they would benefit from being able to use that information to help rapidly determine the assignment. The report in question was assigned the CPT code 88305 by the original coder, so we can be reasonably confident that this was an accurate billing. Because the original coder and our algorithm agree, we are unlikely to reevaluate this case for underbilling. As our models were highly accurate across this corpus, the majority of reports were true positives. For this reason, we have developed methods to automatically display cases of interest (i.e., underbilling candidates).

#### Example of underbilled code identified by the dashboard (CPT 88305 to 88307)

If we restrict the dashboard to only show all false positive predictions, then the dashboard displays to the coder a more focused subset of the reports that all have heightened potential to contain missed CPT codes.

In this example, the case was a distal colon partial resection which was deemed to have been inflamed with reactive epithelial changes. The code originally assigned was CPT code 88305, but the model had revised this to CPT 88307. The example shown in **Figure 3** contains the interpretation plots for assignment of the predicted CPT code 88307. Interpretation plots identified “colon” and “resection” as strong indicators that the report contained the CPT code 88307. While colon biopsies are typically 88305, resections should be 88307 or greater, given that they involve much more pre and post analytic work ^25^. These interpretation plots inform the user of the dashboard on the model’s reasons for assigning 88307, helping them with their assignment. It is also worth pointing out the negative contribution of the word “mucosa.” The algorithm picked up the fact that mucosa examination is not typically associated with codes higher than 88305, and it provides that information to the user as well. In this case, it was not significant enough to change the overall prediction, so the model predicted 88307 despite it.

**Figure 3.**
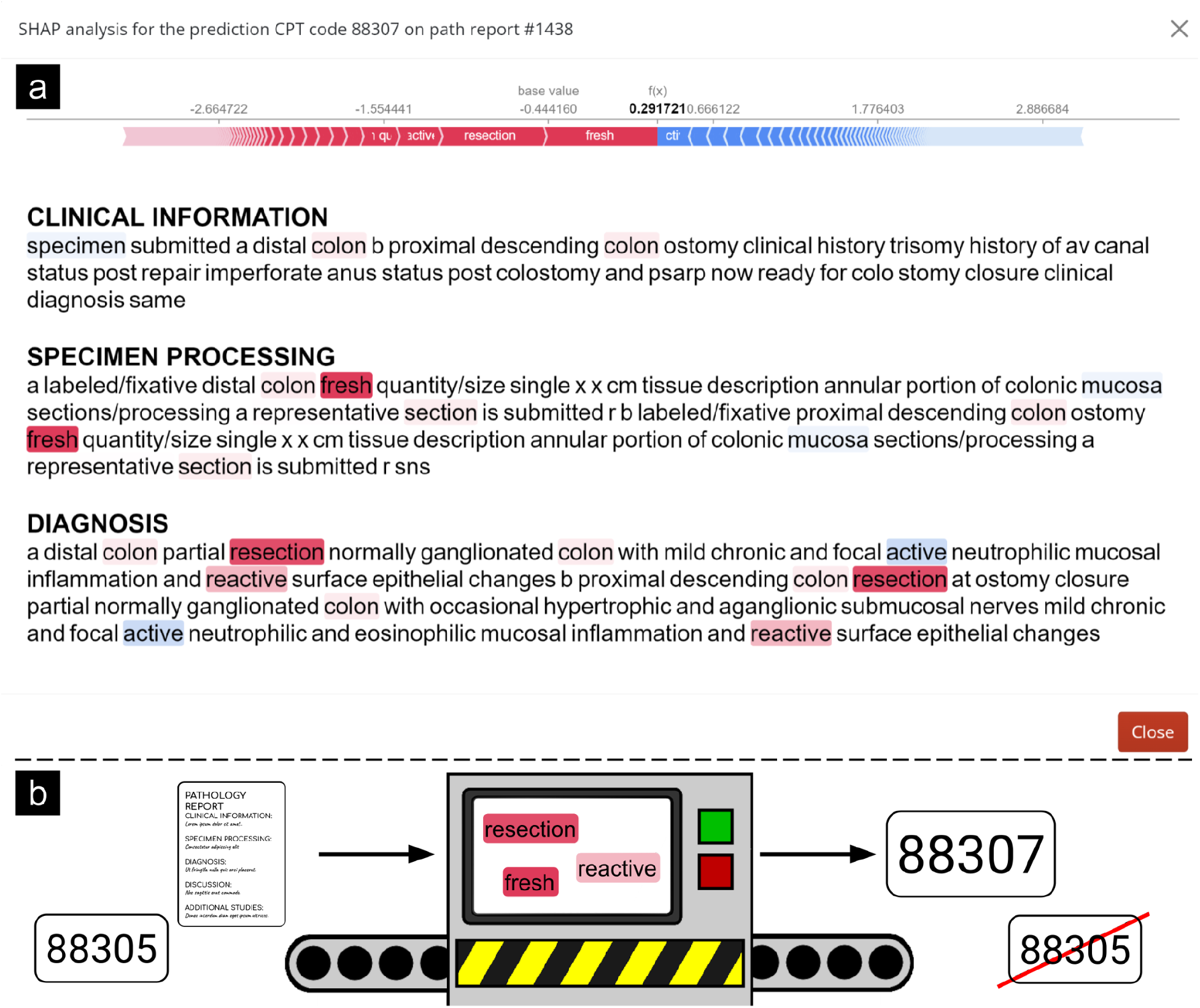
Example interpretation plot for primary CPT code 88307. This report was originally assigned 88305, but our model assigned 88307. The interpretation plot highlighted the words “fresh” and “resection” as important.

If the user wanted more information about why the model chose not to assign 88305, then they could examine the interpretation plots for that code as well **(supplementary material contains the interpretation plots for the class indicating the presence of the code 88305; Supplementary Figures 1-4)**. This interpretation plot shows the new coder exactly why our model disagrees with the original assignment (particularly important for re-assessing underbilled codes, see **Supplementary Figure 5**). In this particular example (**Figure 3**), the model took the presence of “representative” and “resection” to mean that this code was unlikely to represent 88305. The algorithm picked up on the fact that the majority of resections are coded as 88307 or 88309, and it predicted accordingly. The user can use these indicators to rapidly reassess the case, as salient portions of text are highlighted for them. The savings for the hospital associated with this single example would have been in excess of two hundred dollars (**see Methods, “Estimating the Potential Impact of Underbilling”**).

#### Additional examples

In the supplementary material, we have included two additional examples of true positives and two additional examples of false positives / candidate underbilled reports (**Supplementary Figures 6-9**). For instance, **Supplementary Figure 6** displays a report where our model’s assignment and the original coder both assigned the CPT code 88305 and using the SHAP plot correctly identifies esophagus as one of the most informative words (esophagus biopsy falls under criteria for CPT 88305). Similarly, **Supplementary Figure 7** displays a report which demonstrates the true positive assignment of CPT 88307. While examination of other placenta specimen is considered to be a CPT 88305, third trimester pregnancies should be assigned a CPT 88307. This is corroborated using the SHAP functionality in our web application. In **Supplementary Figure 8**, the machine learning model reassigned a CPT 88302 to a CPT 88304. This candidate underbilled case featured an appendectomy as elucidated using SHAP. CPT 88304 is the correct determination because CPT code 88304 does account for several common specimens in which malignancy may rarely present, such as the gallbladder, the appendix and the tonsil. Finally, **Supplementary Figure 9** features inspection of a mass excised from the left palmer hand. A soft tissue mass excised typically warrants assignment of an 88307, save for Lipoma, which is an 88304 ^24^.

### Extrapolating Maximal Underbilling Savings to Future Cohort Assuming Reports were Truly Underbilled

Calculating the price differences for candidate underbilled reports in the study cohort yielded maximal costs savings of $97,084.13 under the assumption that underbill candidates were truly underbilled. Extrapolating these findings to all reports from all pathology subspecialties identified between January 2011 to December 2021, the maximum potential savings could total up to $110,337.38 per year. Of course, while this upper bar sets an estimate of what could be saved if all candidate underbilled reports were in fact underbilled, we expect only a fraction of these reports to be underbilled which will be the subject of future study.

## Discussion

The semi-autonomous assignment of CPT codes will play an important role in billing and revenue for pathology departments which aim to augment their clinical reporting infrastructure. To this end, we have developed a dashboard which can help facilitate the evaluation of these semi-autonomous billing augmentation tools by providing helpful display graphics that inform reassessment. Utilizing interactive digital display dashboards can help further usher in the adoption and embedding of such applications in the electronic health record system pending further validation and user-centered design.

The dashboard featured in this work will allow our team to efficiently review thousands of past cases to provide information on potential revenue lost by the hospital due to underbilling of CPT codes, but it could be used for any task where machine learning is used to interpret free text reports. Overall, model predictions and interpretations from select example cases correlated well with known CPT coding guidelines. This confirmed that the interpretation plots featured in the dashboard could be meaningfully utilized to determine whether the code predicted by the model should replace the original assignment. Every accurate adjustment equates to revenue that our model could have helped save.

## Limitations

There are a few limitations to our study. The model presented in this paper was developed using data at a single hospital and may not be representative of pathology reporting at large. Individuals may write with specific lexical patterns that evolve because of training, peer influence and experience which can complicate analyses and warrant adjustment. XGBoost models stochastically sample predictors, which may not cover the entire vocabulary across the corpus, causing failures in select edge cases. We did not prospectively evaluate the usability of this tool (e.g., execution speed, ergonomics, etc) and plan to do so in a subsequent study. We also plan to evaluate the potential fiscal impact of underbilling on an expanded cohort. There is also considerable clinical information outside of the Pathology report that may prove relevant for case assessment that is unavailable for these approaches. We acknowledge that additional algorithmic finetuning and annotation may improve modeling results. For instance, we assumed that CPT codes used to train the model were taken as ground truth. As it is unlikely our cohort was significantly biased by a few sporadic errors, it should be noted that original codes were likely assigned by a single individual and not re-verified for the purpose of training. We expect the introduced application to rapidly facilitate re-review of mismatches for model training in future iterations of this approach. Furthermore, there may be additional model explanation tools more appropriate than SHAP for medical decision-making tools as SHAP has been critiqued for limited causal interpretability and ethical challenges associated with interpretation in the real-world context (e.g., fairness and stakeholder viewpoints) ^26–30^. This approach may benefit from methods to incorporate expert interpretation. Utilization of these model explanation techniques also requires appropriate education to ensure these tools are appropriately utilized.

## Future Direction

Our next immediate goal is to utilize this application to quantify lost hospital revenue based on model prediction of codes that are higher in complexity than the original assignment (i.e., underbilling). We will make this estimate by comparing the originally assigned codes to the codes that would be assigned with the help of the algorithm when they are flagged as potential false positives in order to quantify how many reports were underbilled. From there we could extrapolate the fiscal burden that missed codes are placing on the hospital ^31–35^.

Additionally, if we find that these models highlight missed codes with high accuracy, that will further enforce the utility of machine learning models for pathology code prediction. As one corrected code could mean the difference of several hundred dollars (e.g., 88309 is priced $450 on average as compared to 88302, which is priced $30 on average), it would not take many to make this correction procedure economically viable. For instance, we found in this study that if underbilling candidates were truly underbilled, this could potentially equate to over $100,000 per year in savings for our institution as extrapolated across an 11-year span, including addendums. This number will be amended/reduced pending review of mismatched cases from local coders to understand the true specificity of this approach though is out of scope for the current work. Should findings prove significant and affirm a consequential estimate, we would therefore want to identify mechanisms to integrate into the coders’ workflow as soon as possible ^16^. Nationally, the average maximum salary for medical coders is $35 per hour ^36–39^. Given that around 1.5% of reports were deemed to be candidate underbilling reports, reexamination of these reports is a worthwhile endeavor versus the amount of time committed (i.e., only one out of every hundred reports would require reexamination). Regardless of the immediate research findings, we will continue to utilize the dashboard to evaluate future, more accurate models (e.g., ones that could potentially improve the coding efficiency of the billing).

We also acknowledge that such a tool could also be configured to evaluate instances of overbilling (e.g., potential fraud), which is especially relevant if there was a systematic finding in a specific specimen type or subspecialty. There are many potential impacts associated with developing algorithms to detect overbilling– for instance, whether detection of such cases leads to loss of revenue or requires additional incentives or additional educational trainings for such a search ^40^. While this is not the specific focus of this research project, utilization of this application can identify general cases of misbilling to improve the ability to “audit” emerging reporting systems and improve their accuracy.

We plan to submit a full research article upon gathering our findings from the analysis of our first models. However, regardless of the algorithm chosen, the dashboard largely serves the same function (i.e., reexamine codes). The basic functions of the dashboard are interoperable to other research domains, and significant changes to improve the functionality are likely not required (while usability improvements will be pursued). The strength of the dashboard is in its ease of use and relative simplicity, allowing researchers to focus more of their time on model development and improvement.

## Conclusion

We have developed an interactive dashboard that can be used to interrogate the assignment of CPT codes via a dashboard agnostic NLP model. The dashboard is additionally model-agnostic, allowing for incorporation of future state-of-the-art approaches. We plan to use the dashboard described in this report to determine the viability of detecting underbilling with our current classification models.

## Data Availability

Data produced in the present study are available upon reasonable request to the authors. We have also included a small online demo at the following URL: https://edit.cpt.code.demo.levylab.host.dartmouth.edu/ (user: edit_user, password: qdp_2022).

## Supplementary

### Supplementary Methods

#### Toggling different SHAP interpretations for CPT code models

This dashboard works primarily with multi-class or multi-target classification of clinical text. Users can toggle between displays that predict the output of *ancillary codes* (i.e., separate models predicting binary endpoints to identify 38 CPT codes, each given a probability 0-1, multiple codes predicted simultaneously) and *primary codes* (i.e., one model to predict presence of one CPT code from a set made up of codes 88302, 88304, 88305, 88307, and 88309). For the *ancillary code* model, the application can only display the SHAP values for one CPT code at a time, so we included a selectable plot of each model’s predictions across the codes in order to quickly switch between the CPT code-specific models. The *primary code* model that we developed also has unique SHAP values for each potential code, so the scatter plot is also used to switch between codes to display correspondent text identified by the model.

#### Filtering documents to focus on false positive reports as candidate underbilling instances

In prediction mode, certain filters can then be applied to select the type of report that the dashboard will present. It is through these filters that we can restrict the dashboard to display false positives only; this is done by selecting the “False predictions” and “Positive only” filters (**Figure 2C**). This filter will show the reports that the model predicts to feature a certain code, but that the original coder did not assign. The coder can then select which codes it identifies from a dropdown menu. Pressing “Next report” updates a CSV file containing the coder’s predictions. From there we can compare their predictions to both the original predictions and our model predictions.

#### Focused search of keywords

Even when not in the hands of coders, the dashboard has a search feature that makes it advantageous to work with. For example, if we believed that our model was particularly good (or perhaps particularly weak) at assigning codes pertaining to examination under a microscope to reports containing the word cytogenetics, then we easily analyze those further. To do this we would select the search by “Report” and search by “Code description” options, and search for “cytogenetics microscope” (**see Figure 2**). Clicking on any of the reports would then display that report on the main dash.

### Supplementary Results

**Supplementary Figure 1:**
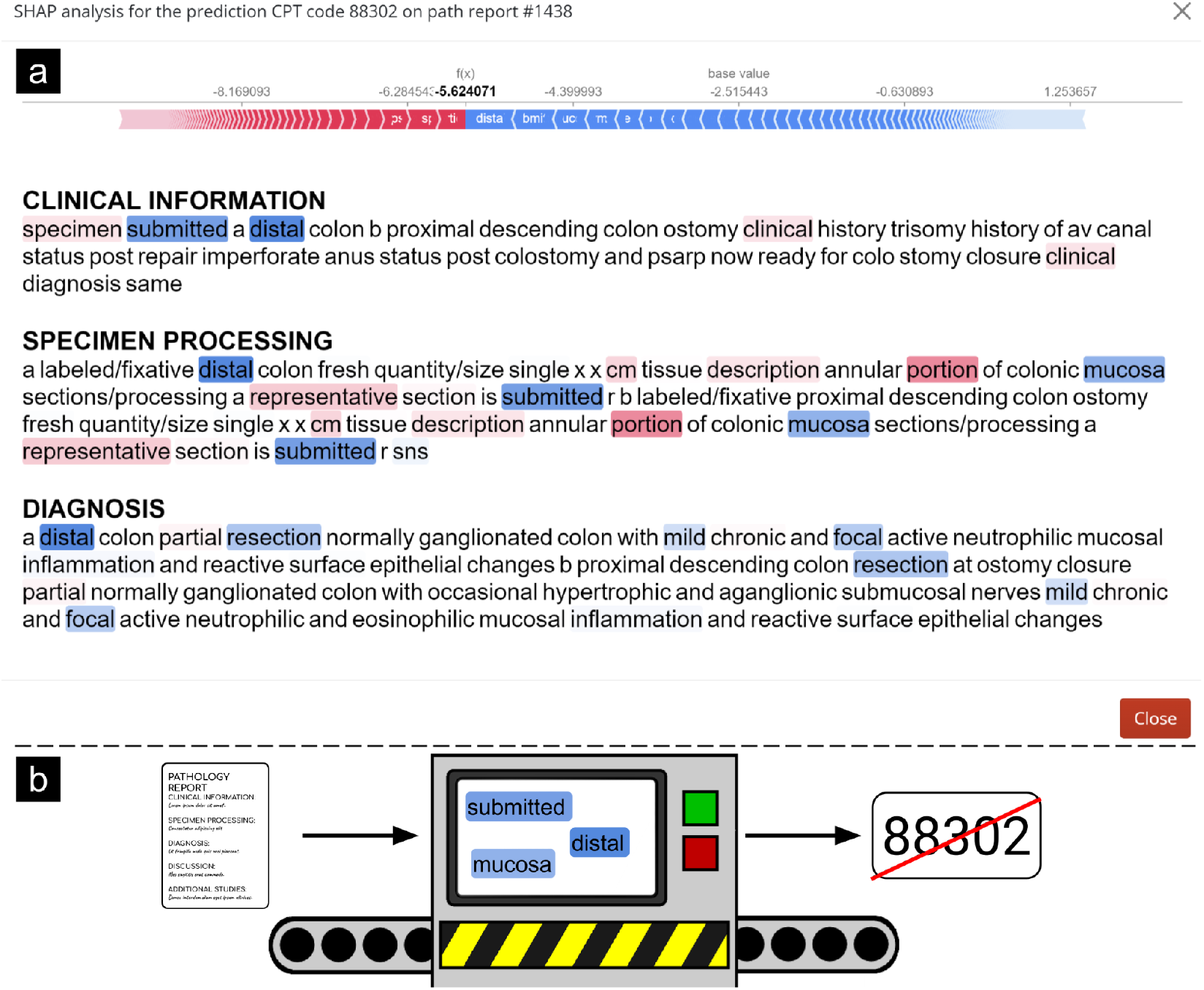
Evaluation of same case from **Figure 3**, depicting words related to CPT 88302 using SHAP on #1438

**Supplementary Figure 2:**
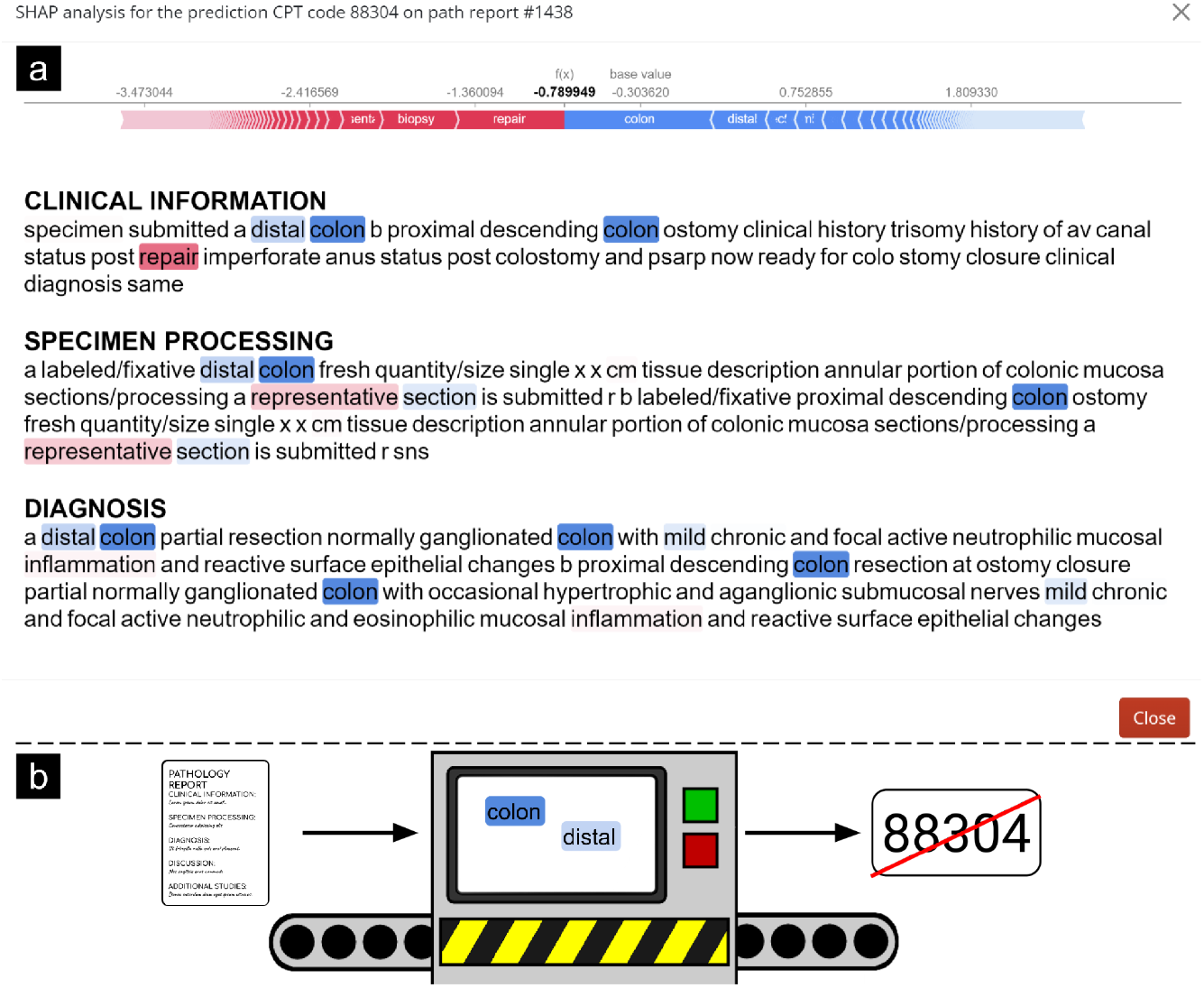
Evaluation of same case from **Figure 3**, depicting words related to CPT 88304 using SHAP on #1438

**Supplementary Figure 3:**
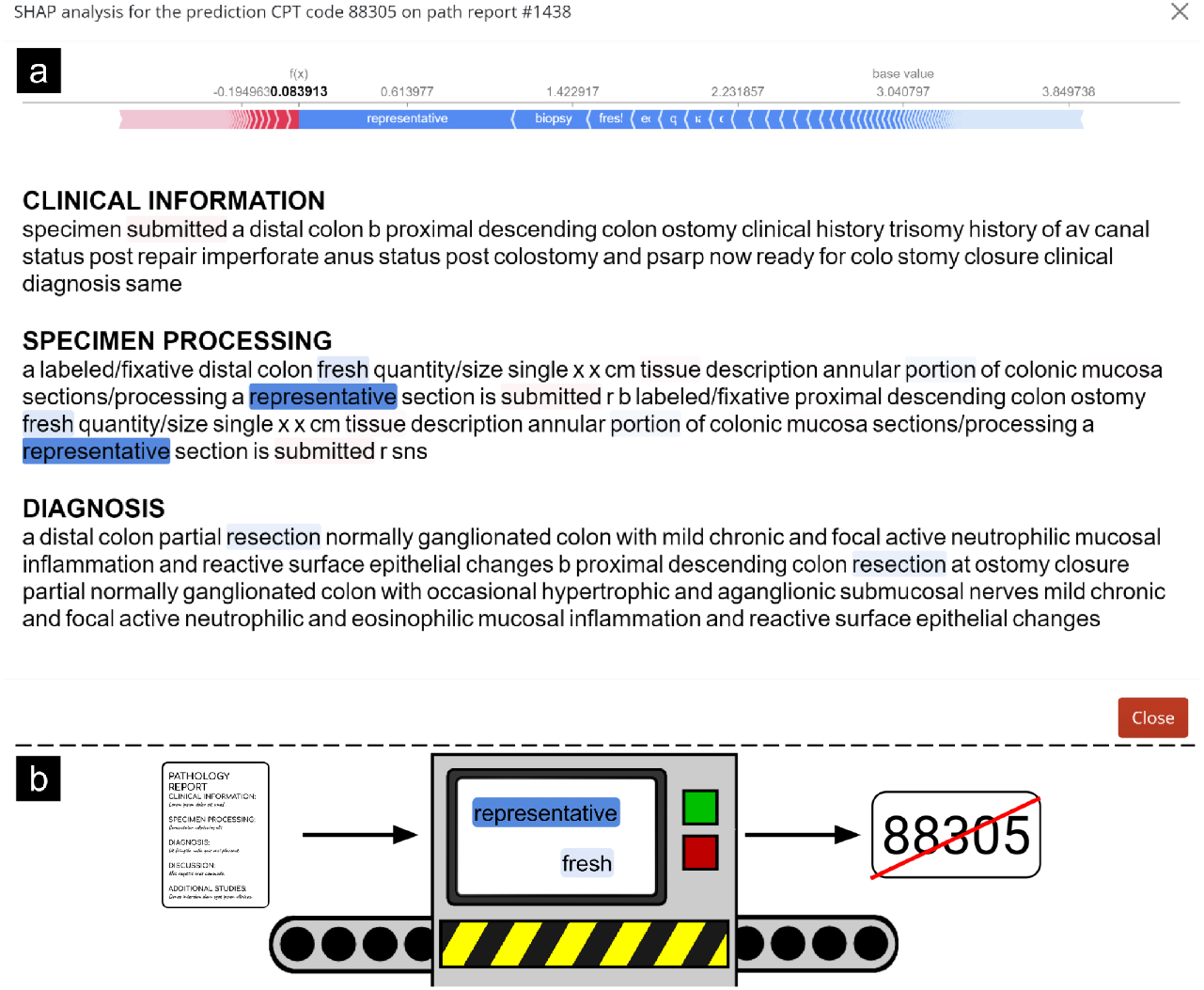
Evaluation of same case from **Figure 3**, depicting words related to CPT 88305 using SHAP on #1438

**Supplementary Figure 4:**
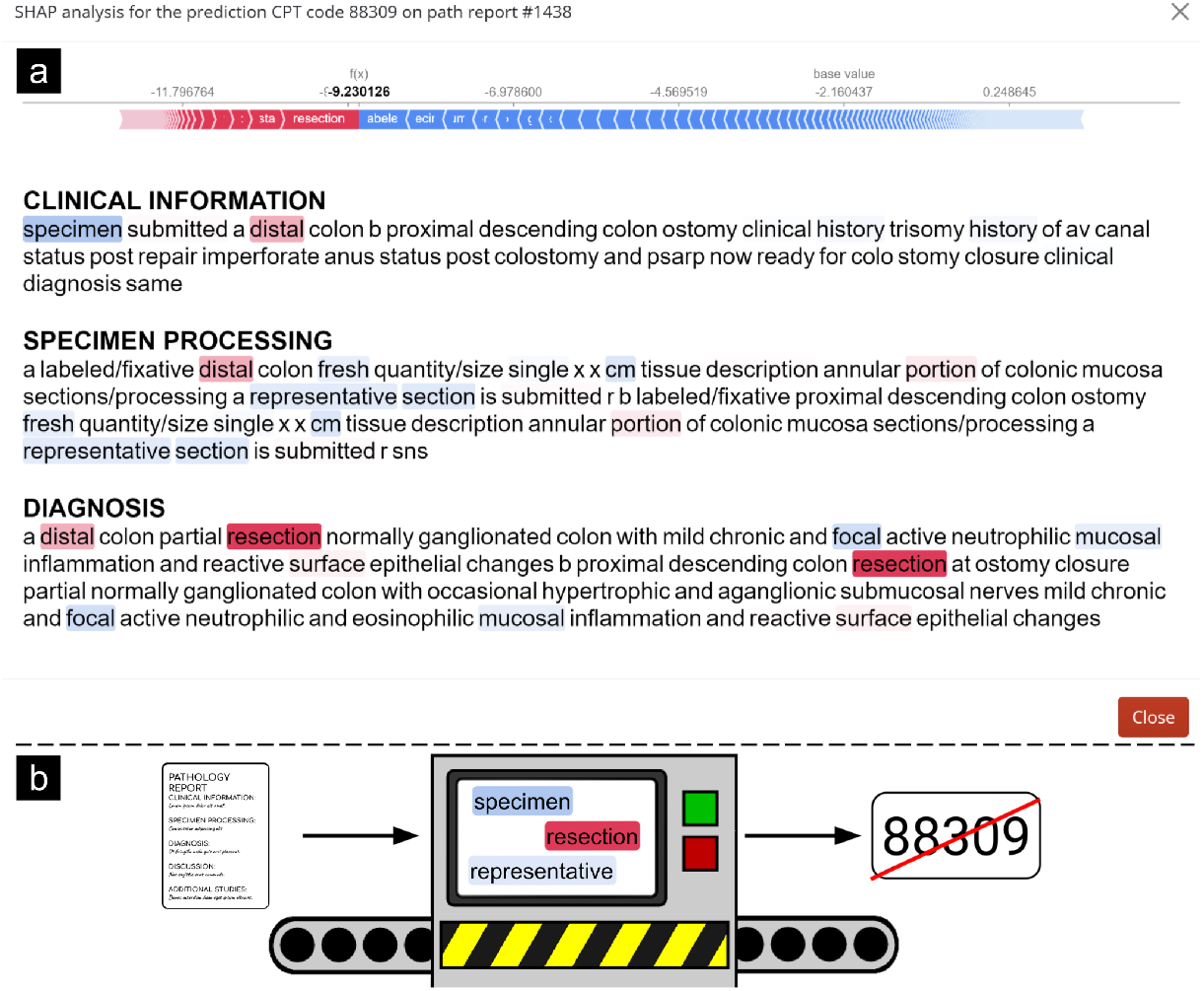
Evaluation of same case from **Figure 3**, depicting words related to CPT 88309 using SHAP on #1438

**Supplementary Figure 5:**
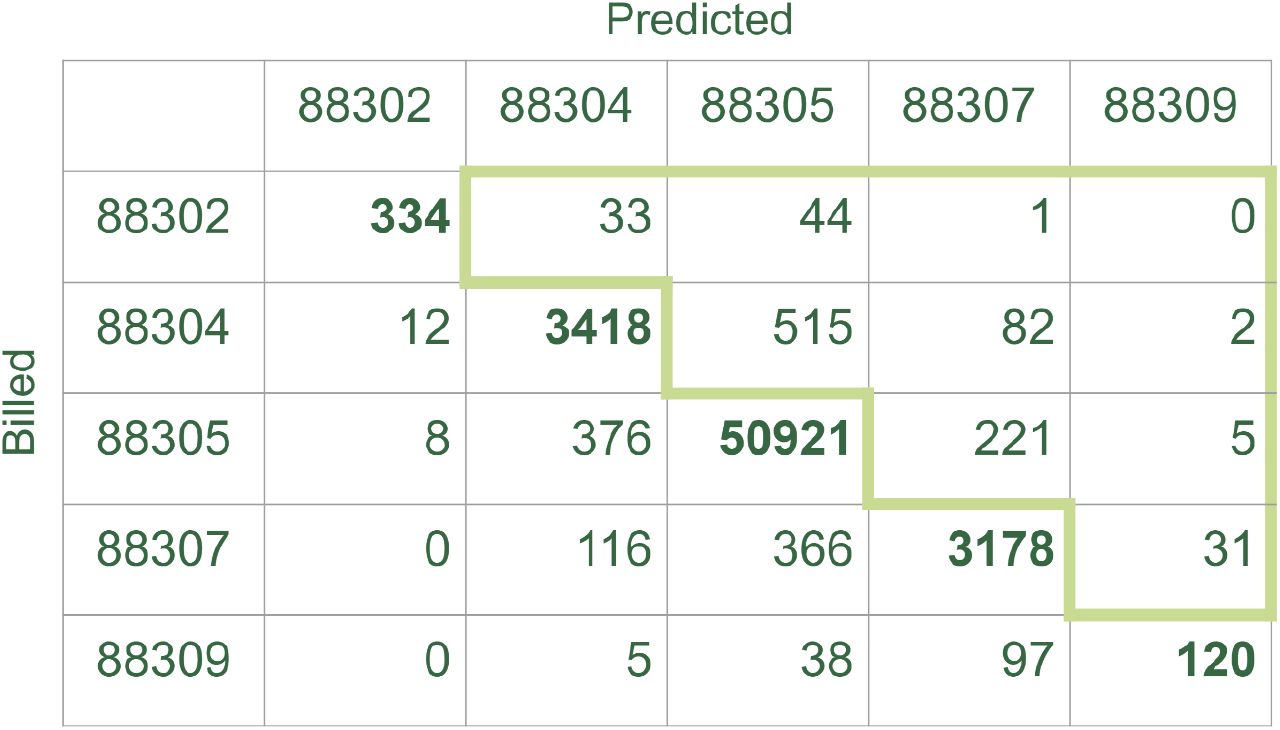
Breakdown of all codes assigned a single primary code in our corpus. Candidate underbilled codes are highlighted in green

**Supplementary Figure 6:**
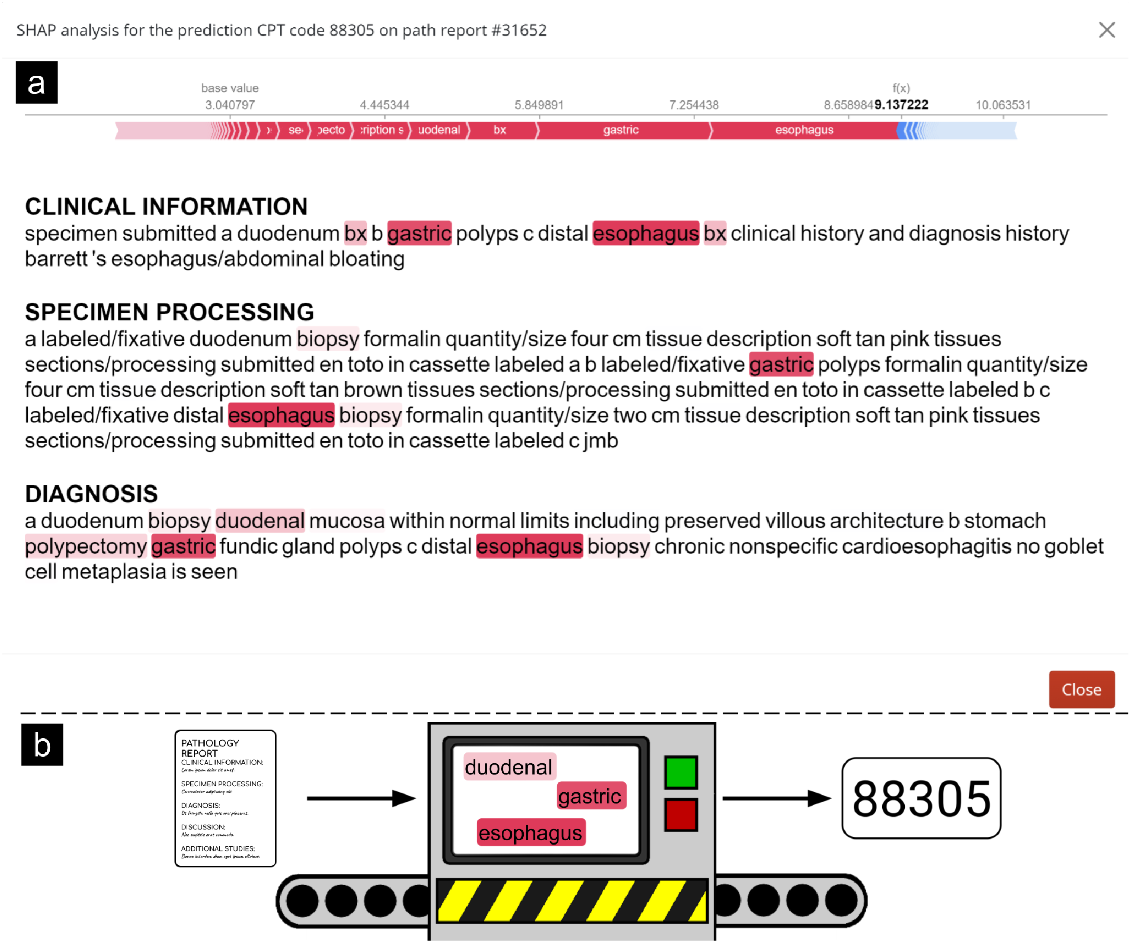
Example interpretation plot for primary CPT code 88305. This report was correctly assigned CPT 88305 by both the coder and model.

**Supplementary Figure 7:**
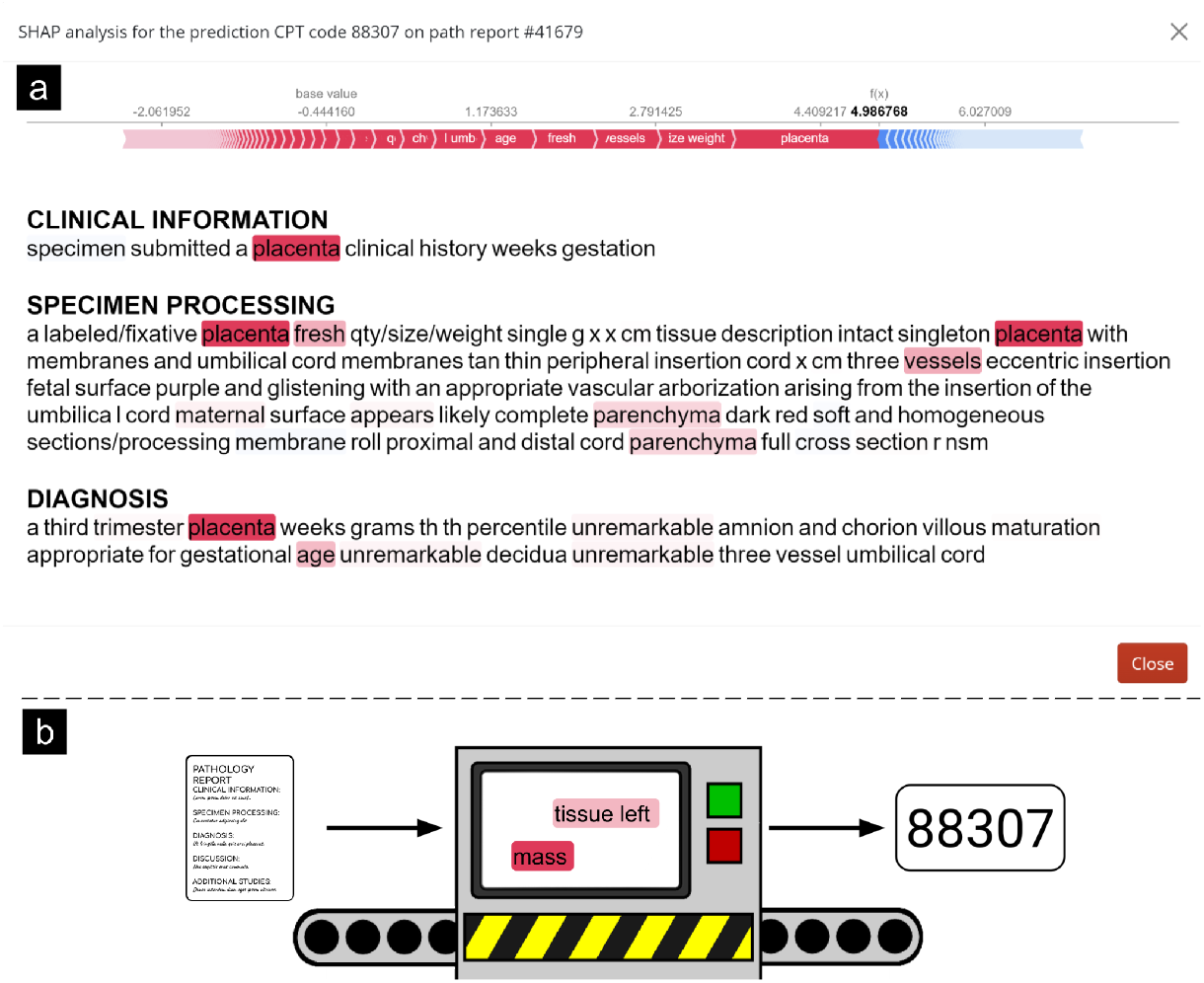
Example interpretation plot for primary CPT code 88307. This report was correctly assigned CPT 88307 by both the coder and model.

**Supplementary Figure 8:**
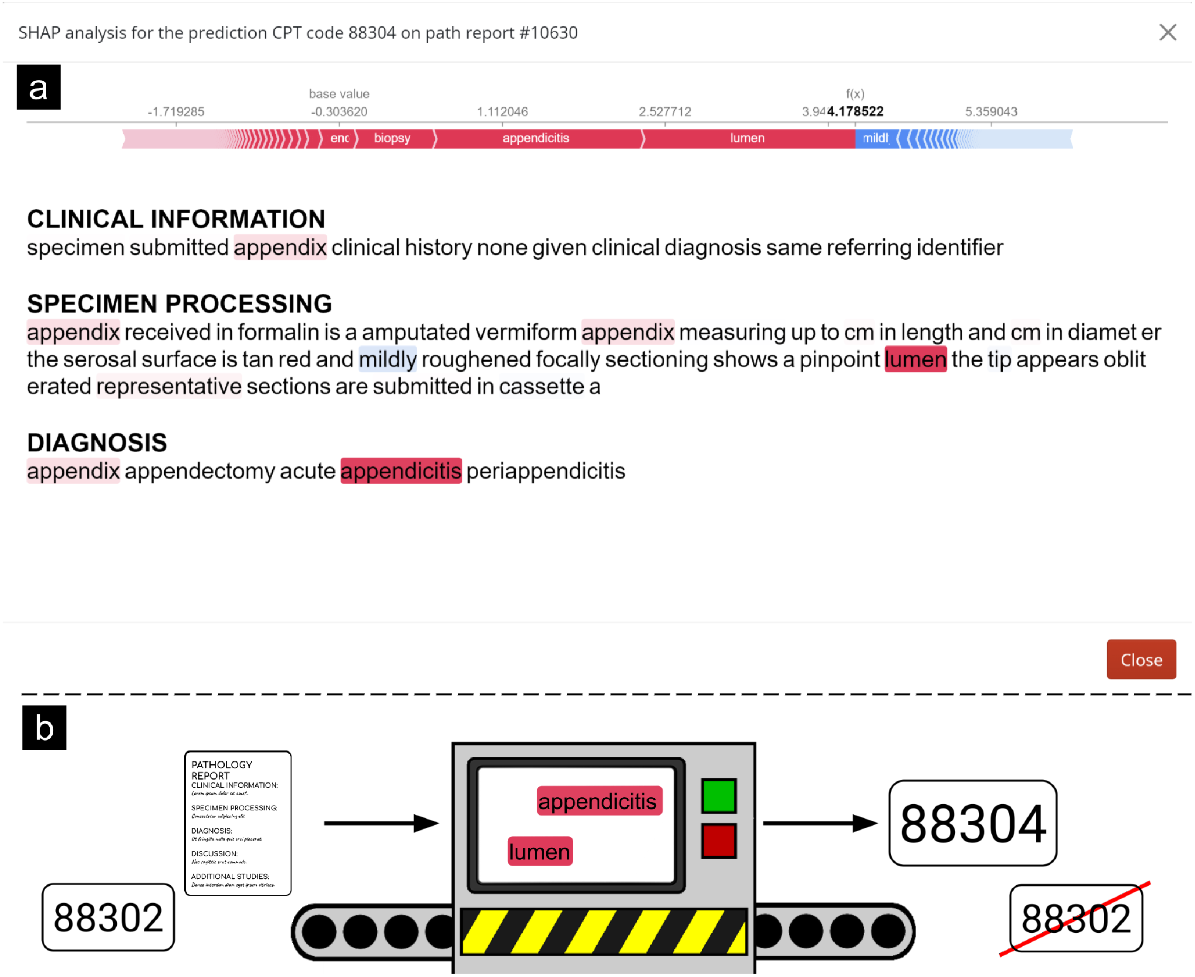
Example interpretation plot for primary CPT code 88304. This report was assigned CPT 88302 by the coder, while the model predicted CPT 88304.

**Supplementary Figure 9:**
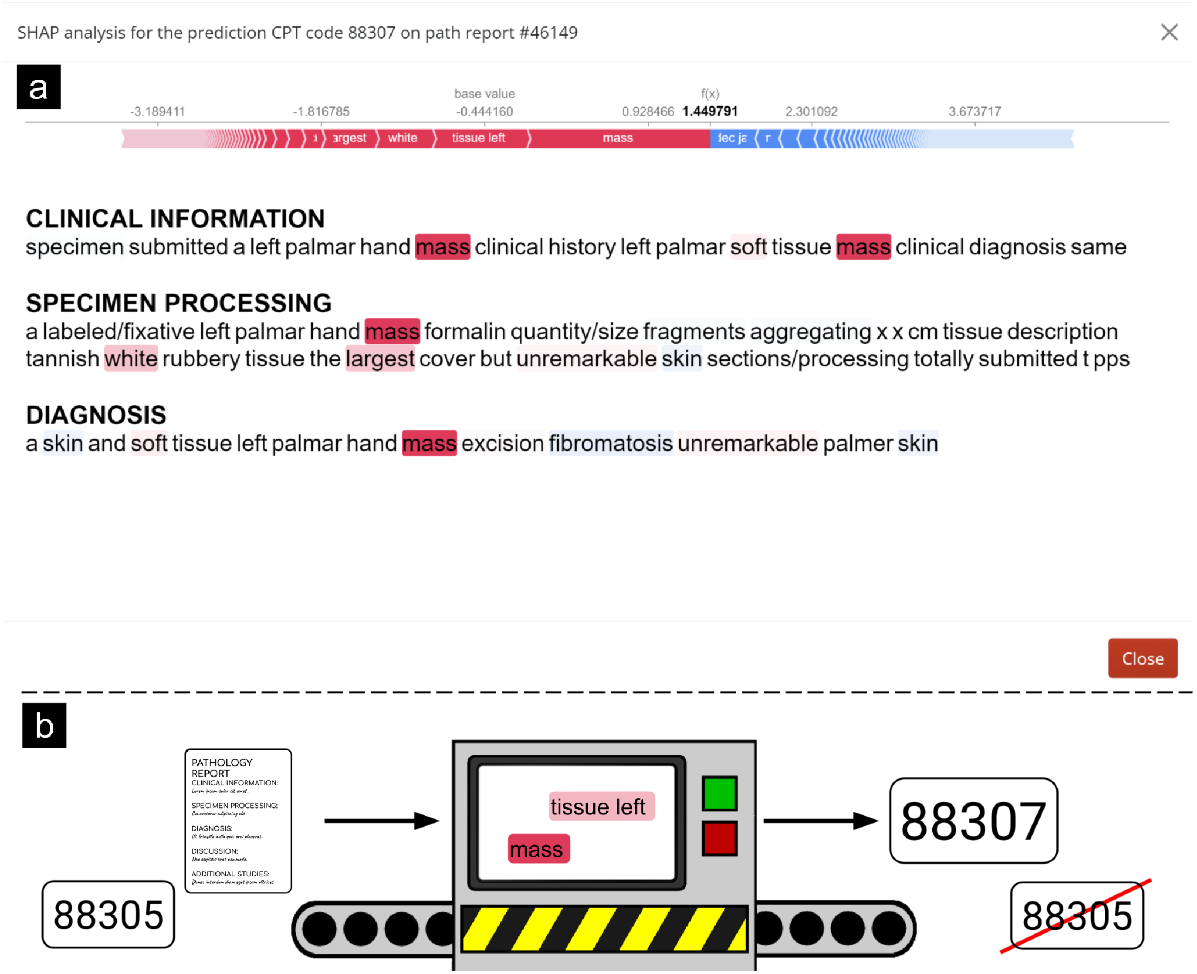
Example interpretation plot for primary CPT code 88307. This report was assigned CPT 88305 by the coder, while the model predicted CPT 88307.

